# Effect of vitamin D on infection and inflammation in patients with cystic fibrosis: a systematic review and meta-analysis

**DOI:** 10.1101/2022.01.03.22268699

**Authors:** Nelufa Begum, Abdullah Al Tarique, Tamara Blake, Dwan Vilcins, Mohammad Zahirul Islam, Nazrul Islam, Robert S Ware, Peter Sly

## Abstract

**Background:** Cystic fibrosis (CF) is a genetic disorder in which the respiratory system gets clogged with mucus leads to progressive lung damage. There is no known cure for CF but several treatments to manage symptoms and reduce complications. Vitamin D deficiency is common in CF associated with increased infection and inflammation. This systematic review and meta-analysis will evaluate the effectiveness of vitamin D treatment in reducing respiratory tract infection and inflammation in patients with CF.

**Methods:** Randomized and quasi-randomised studies in CF patients with control groups will be identified. The antibacterial activity of vitamin D supplementation will help in reducing respiratory tract infection and inflammation in CF. Overall effects of vitamin D in terms of infection and inflammatory markers such as C-reactive protein, inflammatory cytokine interleukin (IL)-6, IL-8, IL-17, IL-23, antimicrobial peptide (LL-37), lung function defined by forced expiratory volume in 1 second (FEV1) %, other assessed respiratory parameters will be calculated using random-effect models. Study quality will be assessed using RoB 2 – A revised Cochrane Risk of Bias tool for randomised trials. The overall quality of evidence for each outcome will be summarised according to the Grading of Recommendations Assessment, Development, and Evaluations (GRADE) framework.

## BACKGROUND

Cystic fibrosis (CF) is a common inherited respiratory disease (1). There is great geographic and ethnic variation in the prevalence of this respiratory disease, but CF has the highest prevalence in Europe, North America, and Australia and remains mainly a disease in Caucasians (2). Patients with CF are more vulnerable to chronic respiratory infection and inflammation in the lower airways, which lead to progressive lung damage. Most patients with CF have reduced capacity to combat chronic respiratory infection, and inflammation leading to respiratory failure is the most common cause of morbidity and mortality in individuals with CF. Although CF cannot be cured, there are several treatment options available to manage symptoms and reduce complications.

Vitamin D deficiency is common in children and adults with CF. Vitamin D is associated with inflammatory response in respiratory tract infections (3-5). Some studies have reported that more than 90% of CF patients have vitamin D deficiency (6, 7), and the correction of vitamin D deficiency reduces inflammation and decreases infection in patients with CF. Vitamin D is a fat-soluble vitamin that is poorly absorbed by individuals with CF unless pancreatic enzyme therapy is adequate. Vitamin D levels are measured routinely at CF annual review, and vitamin supplementation is routine in CF management. To prevent pancreatic insufficiency and deficiency of fat-soluble vitamin D, patients with CF receive age-group-specific vitamin D supplementation according to international CF nutritional guidelines (8). It is well-known that with ongoing supplementation, vitamin D levels improve with pancreatic enzymes (9). A study demonstrated that vitamin D deficiency is associated with alterations in microbiota composition that promote inflammation and supplementation with vitamin D has the potential to impact microbiota composition. It reports that airway microbiota in CF is disrupted by chronic inflammation in the respiratory, and recurrent lung infection (2). Vitamin D deficiency increases susceptibility to infectious diseases and affects control of the inflammation process. Thus, vitamin D supplementation aims to increase vitamin D levels with potential long-term beneficial effects, including antibacterial activity on pulmonary function in patients with CF.

It is well-established that vitamin D deficiency is associated with increased inflammation. Vitamin D has been reported to have an antibacterial effect to reduces respiratory tract infection and inflammation in CF. A study has reported increased vitamin D suppresses the production of proinflammatory cytokines interleukin (IL)-6 and IL-8 and increase the production of the antimicrobial peptide (AMP) and LL-37 from CF respiratory epithelial cells. It indicated that vitamin D was evaluated for its potential to increase LL-37 and reduce inflammation (10).

Vitamin D treatment decreases plasma IL-8 concentration, decreases neutrophil count, reduces inflammation, and improves lung function defined by forced expiratory volume in 1 second (FEV1) % predicted. The study indicated that a high dose of vitamin D improved vitamin D status, which leads a better respiratory function (11). A recent study demonstrated that the anti-inflammatory effect of vitamin D reduced the level of IL-17A and IL-23 in the airway of CF patients with chronic *Pseudomonas aeruginosa* infection and contributed to the exaggerated inflammatory response to pulmonary infection (12).

This review aims to explore whether, in patients with CF, supplementation with vitamin D compared with placebo, is effective in reducing respiratory tract infection and inflammation. Additionally, this review will help identify the relationship between vitamin D status in CF and lung function outcomes.

This review protocol will follow the guidelines for the Preferred Reporting Items for Systematic Reviews and Metanalysis (PRISMA) (13) and is reported here using the Guidance notes for registering a systematic review protocol with PROSPERO provided by the Centre for Reviews and Dissemination (14).

## PROSPERO ITEMS

### 1. Review title

Effect of vitamin D on infection and inflammation in patients with cystic fibrosis: a systematic review and meta-analysis

### 2. Original language title

As above

### 3. Anticipated or actual start date

7 January 2022

### 4. Anticipated completion date

7 July 2022

### 5. Stage of review at time of this submission

Preliminary searches and piloting of the study selection process are underway. After submitting this protocol, the search will be re-run, and eligible journal articles will be identified for inclusion in meta-analyses.

### 6. Named contact

Dr Nelufa Begum

### 7. Named contact email

; Dr Nelufa Begum - Child Health Research Centre, The University of Queensland (uq.edu.au)

### 8. Named contact address

Child Health Research Centre, 62 Graham St, South Brisbane 4101, Qld, Australia

### 9. Named contact phone number

+61 7 3069 7382

### 10. Organisational affiliation of the review

Children’s Health and Environment Program, Child Health Research Centre, The University of Queensland

### 11. Review team members and their organisational affiliations

Nelufa Begum, Children’s Health and Environment Program, Child Health Research Centre, The University of Queensland

Abdullah Al Tarique, Children’s Health and Environment Program, Child Health Research Centre, The University of Queensland

Tamara Blake, Children’s Health and Environment Program, Child Health Research Centre, The University of Queensland

Dwan Vilcins, Children’s Health and Environment Program, Child Health Research Centre, The University of Queensland

Mohammad Zahirul Islam, Children’s Health and Environment Program, Child Health Research Centre, The University of Queensland

Nazrul Islam, Faculty of Health, Queensland University of Technology

Robert S Ware, Menzies Health Institute Queensland, Griffith University

Peter Sly, Children’s Health and Environment Program, Child Health Research Centre, The University of Queensland

## Data Availability

This is a systematic review of public health on cystic fibrosis

## 14. Collaborators

Nil

## 15. Review question

This review aims to explore whether the supplementation of vitamin D is effective in reducing respiratory tract infection and inflammation in patients with cystic fibrosis. We hypothesized that in cystic fibrosis patients who are supplemented with vitamin D, compared with placebo, there will be reduced bacterial infection.

P: Patients with cystic fibrosis

I: Vitamin D treatment/supplementation (any form of doses)

C: Control (placebo) is either no treatment or placebo treatment,

O: infection, inflammation and bacterial killing in the airway.

## 16. Searches

Multiple strategies will be used to identify studies published in English as of December 2021. Sources are four databases - PubMed, EMBASE via Elsevier, CINAHL via EBSCOhost and Web of Science (advanced), from inception to search date. Search strategies for each database were prepared by NB in consultation with other team members and a university library Information specialist. Preliminary searches were conducted using the various suggested terms to guide the study selection process.

## 17. URL to search strategy

The final search strategies as used in this systematic review are detailed in **Appendix I**.

## 18. Condition or domain being studied

Cystic fibrosis

## 19. Participants/population

Children or adult patients with cystic fibrosis will be included in the review. Detailed eligibility criteria are as follows:

The cystic fibrosis patients and

- vitamin D treatment - vitamin D2 or calciferol, vitamin D3, serum25(OH)D, cholecalciferol supplements with any form of doses such as cholecalciferol 1000 IU, ergocalciferol, vitamin complex such as vitamin ABDEK or multivitamin;
- infection and inflammation (sputum or blood microorganisms/pathogens) – macrophage, neutrophil, Streptococci, Pseudomonas aeruginosa, interleukin-6 (IL-6), IL-8, IL-17, IL-23, LL-37, CRP, antimicrobial peptide, etc.

### Exclusion criteria

Cystic fibrosis patients who underwent a lung transplant, UV light therapy for vitamin D will be excluded and no other restrictions will be applied based on comorbidities. Detailed exclusion criteria are as follows:

- No outcomes related to vitamin D treatment
- UV light therapy for vitamin D
- Not in a language that can be translated into English
- Full text not available/conference abstract/poster/meeting abstract/note
- Not original research/case study/review/book chapter/letter/review
- No comparator
- Conference proceedings/editorial/editorial material
- Duplicate study

## 20. Intervention(s), exposure(s)

Any form of vitamin D, say, vitamin D3 (known as cholecalciferol), vitamin D2 (ergocalciferol), etc. at any dose and any duration of intervention. Vitamin D treatment will help to inhibit inflammation and bacterial killing in the airway in cystic fibrosis.

## 21. Comparator(s)/control

Control (placebo) is either no treatment or placebo.

## 22. Types of study to be included

Randomised control trials (RCT) and quasi-randomised control trails such as single centre double-blinded cross-over RCT, multicentre double-blinded RCT, non-blinded RCT. All study designs other than randomized controlled trials (Cohort, Retrospective, case-control, case series, etc,) will be excluded in this review. Studies published as full-text articles that report original research will be eligible for inclusion.

## 23. Context

Studies in patients with cystic fibrosis and vitamin D treatment.

## 24. Main outcome(s)

The effectiveness of vitamin D helps in reducing respiratory tract infection and inflammation in cystic fibrosis patients. Vitamin D treatment will increase serum 25-hydroxyvitamin D (**s**25OHD), decrease plasma IL-8 or IL-23 concentration, which decreases neutrophil count to reduce inflammation in respiratory tract infection. Results from individual studies will be synthesized using random effect models depending on data quality and heterogeneity. For continuous outcomes mean differences (95% confidence intervals) will be calculated, and for categorical outcomes odds ratios with 95% confidence intervals will be calculated to investigate the standardised mean difference (that is, measure of effect size) between the two groups.

### *Measure of effect

No restrictions on the outcome measure of vitamin D. Overall effects of vitamin D in terms of infection and inflammatory on blood or sputum inflammatory biomarkers. All outcomes will be collected at any time frame regardless study length and will analyse separately for short-term (1-16 weeks) and medium term (16-52 weeks) outcomes to gather all possible evidence.

## 25. Additional outcome(s)

Identify the relationship between vitamin D status in CF and lung function outcomes defined by forced expiratory volume in 1 second (FEV1), i.e., respiratory status - FEV1, other assessed respiratory parameters.

## 26. Data extraction (selection and coding)

The bibliographic software, EndNote, will be employed to organize, store, and manage all the references. References retrieved from all four databases will be imported into Endnote. After removal of duplicate references, studies will be selected by three reviewers based on predetermined study inclusion/exclusion criteria. The first selection will be based on title and abstract screening, and the second selection will be based on a full-text screening. Any conflict will be resolved by an independent reviewer. During the full-text screening, any exclusion reasons will be noted to be published in the supplementary material. Data extraction template will be created to gather study information. The template will contain information on author, year of publication, reference, the country in which the study conducted, study type and study design, no. of patients, patient’s allocation, study length, baseline information on (age, s25OHD (ng/ml), other descriptions including lung function – mean (standard deviation)), dose of vitamin D (IU) for intervention and comparator and list of outcomes. NB will extract data from each selected study. The overall quality of evidence for each outcome will assess according to the Grading of Recommendations Assessment, Development, and Evaluations (GRADE) guideline, which classify evidence as either very low, low, moderate, or high.

## 27. Risk of bias (quality) assessment

Two reviewers will perform the study quality assessment independently, with disagreements resolved by a third reviewer. Risk of bias will be assessed using RoB 2 – A revised Cochrane risk-of-bias tool for randomised trials with adjustments made where necessary (15).

## 28. Strategy for data synthesis

Studies included in this review will be grouped according to the primary and secondary outcomes as appropriate. When studies report sufficient data, a meta-analysis will be performed. Results from individual studies will be synthesized using random effect models depending on data quality and heterogeneity. For continuous outcomes mean differences (95% confidence intervals) will be calculated, and for categorical outcomes odds ratios with 95% confidence intervals will be calculated to investigate the standardised mean difference (that is, measure of effect size) between the two groups. Heterogeneity will be measured using I-square statistic. A forest plot will be used to display the summary published findings/results for each study and the amount of study heterogeneity. Publication bias will be tested by generating funnel plots and applying Harbord’s test (16). Analyses will be performed using Stata v17.0 (StataCorp LLC, College Station, TX, USA). The data synthesis will be performed by one reviewer, and the results will be checked by two other reviewers.

## 29. Analysis of subgroups or subsets

Nil

## 30. Type and method of review

Review type - Systematic review, meta-analysis

Health area of the review: Public health on cystic fibrosis

## 31. Language

English

## 32. Country

Australia

## 33. Other registration details

Nil

## 34. Reference and/or URL for published protocol

Protocol will be published on PROSPERO or medRxiv.

## 35. Dissemination plans

It is intended to publish the review article in a peer-reviewed journal. Any modifications to this proposal will be documented in the final published manuscript.

## 37. Details of any existing review of the same topic by the same authors

Nil

## 38. Current review status

Ongoing

## 39. Any additional information

Nil

## 40. Details of final report/publication(s)

### Subject index terms status

Subject indexing assigned by Centre for Reviews and Dissemination (CRD)

### Subject index terms

Cystic Fibrosis, Antibacterial effects of vitamin D; Humans

### Appendix I: Database Search Strategies

#### PubMed

S**earch was performed on November 29, 2021 and returning 620 articles**.

**Query**

(“cystic fibrosis”[tiab] OR CF[tiab] OR “Cystic Fibrosis”[Mesh])

AND

(“vitamin D”[tiab] OR D2[tiab] OR D3[tiab] OR “25-hydroxyvitamin D”[tiab] OR

cholecalciferol*[tiab] OR multivitamin*[tiab] OR

supplementation[tiab] OR supplement*[tiab] OR “Vitamin D”[Mesh])

AND

(Antibacterial[tiab] OR Anti-bacterial[tiab] OR antimicrobial[tiab] OR anti-microbial[tiab]

OR infection*[tiab] OR “respiratory infection*”[tiab] OR “respiratory bacteria”[tiab] OR

inflammat*[tiab] OR pro-inflammat*[tiab] OR CRP[tiab] OR “c-reactive protein”[tiab] OR

phagocytosis[tiab] OR macrophage*[tiab] OR neutrophil*[tiab] OR NE[tiab] OR

streptococc*[tiab] OR “airway surface liquid”[tiab] OR ASL[tiab] OR “Pseudomonas

aeruginosa”[tiab] OR “antimicrobial peptide”[tiab] OR IL[tiab] OR interleukin*[tiab] OR IL-

6[tiab] OR IL-8[tiab] OR IL-17[tiab] OR IL-23[tiab] OR LL-37[tiab] OR cytokin*[tiab] OR

microbiota[tiab] OR microbiome[tiab] OR “Anti-Infective Agents”[Mesh] OR

“Inflammation”[Mesh] OR “Infections”[Mesh])

#### EMBASE (via Elsevier)

S**earch was performed on November 29, 2021 and returning 2**,**010 articles**.

**Query**

(“cystic fibrosis”:ti,ab OR CF:ti,ab OR “Cystic Fibrosis”/exp)

AND

(“vitamin D”:ti,ab OR D2:ti,ab OR D3:ti,ab OR “25-hydroxyvitamin D”:ti,ab OR

cholecalciferol*:ti,ab OR multivitamin*:ti,ab OR supplementation:ti,ab OR supplement*:ti,ab

OR “Vitamin D”/exp)

AND

(Antibacterial:ti,ab OR Anti-bacterial:ti,ab OR antimicrobial:ti,ab OR anti-microbial:ti,ab OR

infection*:ti,ab OR “respiratory infection*”:ti,ab OR “respiratory bacteria”:ti,ab OR

inflammat*:ti,ab OR pro-inflammat*:ti,ab OR CRP:ti,ab OR “c-reactive protein”:ti,ab OR

phagocytosis:ti,ab OR macrophage*:ti,ab OR neutrophil*:ti,ab OR NE:ti,ab OR

streptococc*:ti,ab OR “airway surface liquid”:ti,ab OR ASL:ti,ab OR “Pseudomonas

aeruginosa”:ti,ab OR “antimicrobial peptide”:ti,ab OR IL:ti,ab OR interleukin*:ti,ab OR IL-

6:ti,ab OR IL-8:ti,ab OR IL-17:ti,ab OR IL-23:ti,ab OR LL-37:ti,ab OR cytokin*:ti,ab OR microbiota:ti,ab OR

microbiome:ti,ab OR “AntiInfective Agent”/exp OR Inflammation/exp

OR Infection/exp)

AND

[embase]/lim

#### CINAHL (via EBSCOhost)

S**earch was performed on November 29, 2021 and returning 121 articles**.

**Query**

((TI “cystic fibrosis” OR AB “cystic fibrosis”) OR (TI “CF” OR AB “CF”) OR (MH “Cystic Fibrosis”+))

AND

((TI “vitamin D” OR AB “vitamin

D”) OR (TI D2 OR AB D2) OR (TI D3 OR AB D3) OR (TI “25-hydroxyvitamin

D” OR AB “25-hydroxyvitamin

D”) OR (TI cholecalciferol* OR AB cholecalciferol*) OR (TI multivitamin* OR AB multivit amin*) OR (TI supplementation OR AB supplementation) OR (TI supplement* OR AB suppl ement*) OR (MH “Vitamin D”+))

AND

((TI Antibacterial OR AB Antibacterial) OR (TI Anti-bacterial OR AB Anti-

bacterial) OR (TI antimicrobial OR AB antimicrobial) OR (TI anti-microbial OR AB anti-

microbial) OR (TI infection* OR AB infection*) OR (TI “respiratory

infection*” OR AB “respiratory infection*”) OR (TI “respiratory

bacteria” OR AB “respiratory bacteria”) OR (TI inflammat* OR AB inflammat*) OR (TI pro-

inflammat* OR AB pro-inflammat*) OR (TI CRP OR AB CRP) OR (TI “c-reactive

protein” OR AB “c-reactive

protein”) OR (TI phagocytosis OR AB phagocytosis) OR (TI macrophage* OR AB macropha

ge*) OR (TI neutrophil* OR AB neutrophil*) OR (TI NE OR AB NE) OR (TI streptococc*

OR AB streptococc*) OR (TI “airway surface liquid” OR AB “airway surface

liquid”) OR (TI ASL OR AB ASL) OR (TI “Pseudomonas

aeruginosa” OR AB “Pseudomonas aeruginosa”) OR (TI “antimicrobial

peptide” OR AB “antimicrobial

peptide”) OR (TI IL* OR AB IL*) OR (TI interleukin* OR AB interleukin*) OR (TI IL-

6 OR AB IL-6) OR (TI IL-8 OR AB IL-8) OR (TI IL-17 OR AB IL-17) OR (TI IL-

23 OR AB IL-23) OR (TI LL-37 OR AB LL-

37) OR (TI cytokin* OR AB cytokin*) OR (TI microbiota OR AB microbiota) OR (TI micro

biome OR AB microbiome) OR (MH “Antiinfective

Agent”+) OR (MH Inflammation+) OR (MH Infection+))

#### Web of Science (advanced)

S**earch was performed on November 29, 2021 and returning 751 articles**.

**Query**

((TI=“cystic fibrosis” OR AB=“cystic fibrosis”) OR (TI=CF OR AB=CF) OR ALL=“Cystic Fibrosis”)

AND

((TI=“vitamin D” OR AB=“vitamin

D”) OR (TI=D2 OR AB=D2) OR (TI=D3 OR AB=D3) OR (TI=“25-hydroxyvitamin

D” OR AB=“25-hydroxyvitamin

D”) OR (TI=cholecalciferol* OR AB=cholecalciferol*) OR (TI=multivitamin* OR AB=multi vitamin*) OR (TI=supplementation OR AB=supplementation) OR (TI=supplement* OR AB=supplement*) OR ALL=“Vitamin D”)

AND

((TI=Antibacterial OR AB=Antibacterial) OR (TI=Anti-bacterial OR AB=Anti-

bacterial) OR (TI=antimicrobial OR AB=antimicrobial) OR (TI=anti-microbial OR AB=anti-

microbial) OR (TI=infection* OR AB=infection*) OR (TI=“respiratory

infection*” OR AB=“respiratory infection*”) OR (TI=“respiratory

bacteria” OR AB=“respiratory

bacteria”) OR (TI=inflammat* OR AB=inflammat*) OR (TI=pro-inflammat* OR AB=pro-

inflammat*) OR (TI=CRP OR AB=CRP) OR (TI=“c-reactive protein” OR AB=“c-reactive

protein”) OR (TI=phagocytosis OR AB=phagocytosis) OR (TI=macrophage* OR AB=macro

phage*) OR (TI=neutrophil* OR AB=neutrophil*) OR (TI=NE OR AB=NE) OR (TI=strepto

cocc* OR AB=streptococc*) OR (TI=“airway surface

liquid” OR AB=“airway surface liquid”) OR (TI=ASL OR AB=ASL) OR (TI=“Pseudomonas

aeruginosa” OR AB=“Pseudomonas aeruginosa”) OR (TI=“antimicrobial

peptide” OR AB=“antimicrobial

peptide”) OR (TI=IL OR AB=IL) OR (TI=interleukin* OR AB=interleukin*) OR (TI=IL-

6 OR AB=IL-6) OR (TI=IL-8 OR AB=IL-8) OR (TI=IL-17 OR AB=IL-17) OR (TI=IL-

23 OR AB=IL-23) OR (TI=LL-37 OR AB=LL-

37) OR (TI=cytokin* OR AB=cytokin*) OR (TI=microbiota OR AB=microbiota) OR (TI=microbiome OR AB=microbiome) OR ALL=“Anti-Infective

Agents” OR ALL=Inflammation OR ALL=Infections)

